# Delay-related harm: direct and indirect impacts of boarding medical patients in the Emergency Department on the urgent and emergency care pathway. A retrospective observational cohort study

**DOI:** 10.1101/2025.02.21.25322666

**Authors:** Nicholas C Howlett, James A Cameron, Richard M Wood

**Author notes:** Correspondence: Dr James Cameron.

## Abstract

**Background:** Previous studies have indicated that crowding within the Emergency Department (ED) is associated with longer lengths of stay in the ED and higher mortality. Boarding, the time patients spend waiting for an inpatient bed after ED assessment, represents a clinically unproductive delay, and occupies scarce ED resources. We aimed to explore the impact of medical patients boarding not only on their outcomes but also indirectly on other patients in the ED and in and awaiting ambulances.

**Methods:** A retrospective cohort study using routine data for 3 EDs in England from June 2021 to May 2024 was performed. Direct outcomes of medical patient boarding time were investigated: inpatient length of stay, 30-day readmission rate and mortality. Indirect outcomes of medical patient boarding levels consisted of time in ED for non-admitted patients, ambulance handover times, and ambulance response times. Regression analysis was used to model each relationship while controlling for other potentially confounding variables.

**Results:** In all, data on 223,856 ambulance responses, 117,800 ambulance handovers, 367,985 non-admitted ED patients, and 46,976 medical admissions were studied. Medical patients, covering two-thirds of ED admissions, constituted 82% of total ED boarding time. Regression analysis showed that for a typical 25-bed ED, each additional five medical boarders was associated with an extra 12 and 39 minutes for Category 2 and 3 ambulance response times (p<0.001) and an extra 20 minutes for ambulance handover times (p<0.001). For admitted medical patients, each additional 4 hours of boarding time was associated with an extra 13 hours inpatient length of stay (p<0.001) and a 6% increase in odds of 30-day mortality (p<0.01).

**Conclusion:** Boarding of medical patients in the ED is associated with direct harm for those patients, and indirect harms for other patients in the ED and awaiting ambulances.

**What is already known on this topic:** A prolonged length of stay in the Emergency Department is known to increase mortality for those patients.

**What this study adds:** Medical admissions make up 85% of boarding time in the ED waiting for an inpatient bed. This delay affects them directly with increased mortality, readmission and length of stay. It affects other patients with delayed ambulance offload and ambulance response times for other patients.

**How this study might affect research, practice or policy:** The boarding delay for medical patients in the ED results in a longer inpatient length of stay and therefore fewer beds available for future patients. Stopping this practice would free up much needed hospital capacity, improve ambulance response times, and reduce mortality.

## 1. Introduction

Crowding of Emergency Departments (EDs) is a significant problem in many countries [1,2]. Causes may be broadly classified to: input, driven by too many (complex) patient arrivals; throughput, constrained by staffing numbers, diagnostics capacity, beds and productivity; and output, limited by a lack of inpatient beds for patients requiring admission, which, in turn, is typically driven by insufficient intermediate and social care capacity [3,4]. Consequences of ED crowding include delayed ambulance handover times [5] and longer ED length of stay (LOS), which, in turn, associates with increased mortality [6-9] and elevated subsequent inpatient LOS [5,10,11].

Prior to admission, patients in ED receive an initial assessment, investigations and treatment before a decision to admit is made, this should be well within the national 4-hour target. Following the decision to admit, any further time spent in the ED – referred to as ‘boarding’ – represents an unproductive delay. When demand for ED beds exceeds capacity, these patients are often positioned in corridors or other escalation areas. Not only are these environments inferior insofar as clinical care, but such care diverts resources from other patients who still require treatment in ED.

Medical patients (those requiring admission under the Medical team rather than e.g. Surgical or Orthopaedic) are typically the largest cohort requiring admission and experience the longest waits for an inpatient bed, and so form the majority of patients boarding in the ED [12,13]. Our study seeks to understand and quantify the impacts of medical patient boarding on six indicators of patient harm, or Outcome Measures (OMs), covering ED input, throughput and output. The hypothesis is that the boarding of medical patients causes harm indirectly to other patients as well as directly to those experiencing boarding themselves.

## 2. Methods

### 2.1 Study setting and data sources

This study was conducted within the Bristol, North Somerset and South Gloucestershire (BNSSG) Integrated Care System (ICS) in South West England. BNSSG serves a one-million-person population across diverse communities and has a similar age profile to England. The region includes three hospitals with EDs: Southmead Hospital (20 beds including 6 resuscitation beds), Bristol Royal Infirmary (24 beds including 8 resuscitation beds), and Weston General Hospital (13 beds including 4 resuscitation beds).

Data on ambulance response and handover times was sourced from the Ambulance Data Set [14]. ED data was sourced from the Emergency Care Data Set [15]. Admitted patient LOS and comorbidity measures were sourced from the Secondary Uses Service (SUS) Admitted Patient Care (APC) dataset, a derivative of the Hospital Episode Statistics (HES) dataset [16]. Mortality data was sourced from the Civil Registrations of Death dataset [17]. Where applicable, the data sources are linked via a pseudonymised patient ID and event time stamps. All data were anonymised with no patient identifiable information included and all necessary permissions were obtained for use of this data in the study.

### 2.2 Study design and statistical approach

The link between the OMs and harm (and therefore the rationale for selecting the OMs) is as follows. For the three considered indirect indicators of patient harm, ‘upstream’ of ED boarding, we include ambulance response time (OM1) and ambulance handover time (OM2), which are both known to associate with poorer patient outcomes including mortality [18-20], and time until ED treatment for non-admitted patients (OM3), given the link between time to ED treatment and mortality [21]. For the three considered direct indicators of patient harm, ‘downstream’ of ED boarding, we consider inpatient LOS (OP4), which has been found to associate with poor patient outcomes [22], as well as readmission (OM5) and mortality (OM6) within 30 days of admission from ED.

The aim of this study was approached through a retrospective observational cohort analysis in which the six considered Outcome Measures (OM1-6) were examined separately. In each such instance, the approach involved statistically modelling the level of association between the relevant boarding-related ‘exposure’ and the OM, while controlling for the effect of other possibly confounding variables on the OM. In all cases, regression models were fit, with use of a linear regression for continuous-valued OMs (OM1-4) and a logistic regression for binary-valued OMs (OM5-6). The STROBE checklist was followed within this study.

In assessing for indirect, or ‘upstream’, impacts of medical patient boarding (OM1-3), the cohorts were defined by those who, between 1 June 2021 and 1 June 2024, experienced the following OM-related events: ambulance dispatched (OM1); conveyed to ED by ambulance (OM2); attended ED but not admitted (OM3). Ambulance response time (OM1) was partitioned into three groups according to the (decreasing) severity level assigned to the call: Category 1 (OM1a); Category 2 (OM1b); and Category 3 (OM1c). Confounding variables consisted of corresponding patient age (discretised to decades, to account for possible non-linearity), sex, deprivation decile (per Index of Multiple Deprivation), time of event (to 6-hour bands), month of event, whether the event occurred on a weekend, and the ED occupancy at the time of event excluding medical boarders (so isolating the impact of medical boarders from overall ED occupancy). The exposure was defined by the number of medical boarders at the ED at the time of the event. This was standardised to account for ED size, by dividing the number of boarders by permanent bedded capacity (including resuscitation beds).

In assessing for direct, or ‘downstream’, impacts of medical patient boarding (OM4-6), the single cohort was defined by those who had been admitted to an inpatient ward from ED to specialties General Internal Medicine, Acute Internal Medicine, and Geriatric Medicine between 1 June 2021 and 2 May 2024 (i.e., leaving a 30-day period for readmission and mortality per OM5-6), including only the first admission for each patient within the aforementioned period and including only admissions where all confounding variables were present. Confounding variables consisted of corresponding patient age, sex, deprivation decile, time of admission, month of admission, whether the admission occurred on a weekend, the patient’s comorbidity score (as calculated by the Van Walraven Elixhauser measure [23] using ICD-10 codes from the admission), the number of ED attendances within the past 60 days before the admission (excluding the attendance directly before the admission), and the 4-hour ED performance (proportion of ED conclusions within 4 hours) at the time of admission (so isolating the impact of medical boarding time from overall ED performance). Squared terms were included for the three continuous-valued confounders: comorbidity score, number of ED attendances within the past 60 days, and the 4-hour ED performance at the time of admission. The exposure was defined by medical boarding time at the ED.

From the resulting fitted regression models for OM1-6, the effect size and accompanying p-values were output. To ease interpretability, the effect size on the indirect/upstream impacts (OM1-3) were calculated on the basis of a 20% increase in the exposure (number of medical boarders) and the effect size on the direct/downstream impacts (OM4-6) were calculated based on a 4-hour increase in the exposure (medical boarding time). Full details for all regression model parameters, including raw effect sizes (beta and odds ratio respectively for the linear and logistic regressions), can be found in the Supplementary Material.

Additionally, for the indirect/upstream impacts (OM1-3), the linear regressions were refitted while excluding the exposure variable (number of medical boarders). For each OM, the models were used to produce estimates of the OM for each patient in the cohort, and residuals were calculated, to which the cohort-level empirical means were added. This aimed to provide standardised versions of the observations with the effects of the confounders removed. Extracting the corresponding number of medical boarders at the respective event times – expressed as a percentage of ED bedded capacity and binned to 22 5%-width bands from 0% to 110% (this range encompassed 95% of the overall data) – boxplots were produced, showing the range of OM values for each of these 22 standardised exposure bins. This facilitated a richer understanding of the relationship between the exposure variable and the OM, such as in assessing potential areas of non-linearity. It also allowed for the inclusion of national performance targets, in contextualising the results. These included: 90% of Category 1, 2 and 3 ambulance response times within 15, 40, and 120 minutes respectively (OM1) [24]; 90% of ambulance handover times within 30 minutes (OM2) [25]; and 78% of ED patients seen for treatment within 4 hours (OM3) [26].

For the direct/downstream impacts (OM4-6), no such targets existed and, moreover, there was insufficient numbers of event outcomes in OM5-6 to take a similar approach. Instead, the associations between the exposure variable (medical boarding time) and OMs were illustrated through predictor effect displays [27], in which the exposure is varied while holding constant the confounding variables at typical values (mean for continuous variables and proportions for categorical variables)

## 3. Results

The mean time to decision to admit under Medicine across the three hospitals was 190 minutes. Medical patients constituted two-thirds of ED admissions, had substantially longer boarding time (mean 384 vs 164 minutes), and so represented a large (82%) proportion of total admission delay time.

Table 1 presents a descriptive summary of the cohorts used in this study, in terms of the different events which constitute each cohort and their respective values for the relevant exposure variable and confounders at the time of the event. These results are presented after the exclusion of any events with missing data for any confounding variables. Data completeness varied across all OMs. Missing data were predominantly observed for deprivation decile, with a small number of instances of missingness in patient sex. In total 34,726 events were excluded from OM1 (15.5% of all cases for this OM), 13,633 events were excluded from OM2 (11.5%), 10,065 events were excluded from OM3 (2.7%), and 1,602 events were excluded from OMs 4-6 (3.5%).

**Table 1.**
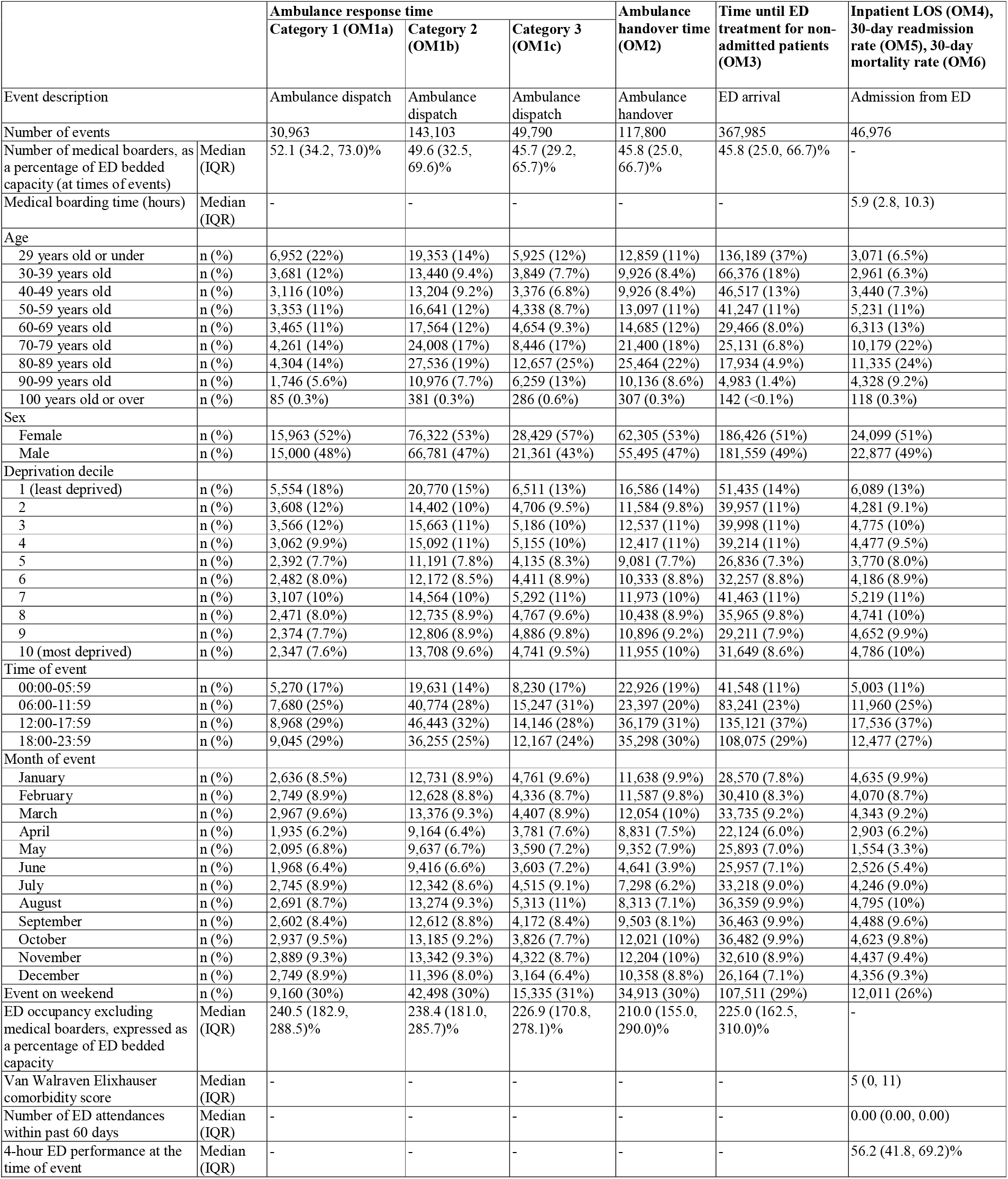
Descriptive summary of the cohorts used to investigate the exposure effect on the various considered Outcome Measures (OMs).

The main results of this study are summarised in Table 2, by presentation of the effect size (and corresponding p-value) of the boarding-related exposures on each OM1-6. In general, the ‘direction’ of the associations is consistent with an adverse patient outcome across all OMs, with high levels of statistical evidence (p<0.05) demonstrated, especially for OM1-4 (p<0.001).

**Table 2.** Effect size, detailing the impact of changes to the exposure variables on the respective Outcome Measures (OMs), including 95% confidence intervals (CIs). P-values indicate the level of statistical evidence in the associations.

| Exposure | Outcome | Effect size (95% CI) | P-value |
| --- | --- | --- | --- |
| + 20% increase in the number of medical boarders, as a percentage of ED bedded capacity (e.g., an extra 5 medical boarders for a 25-bed ED) | Ambulance response time Category 1 (OM1a) | 0.5 (0.4, 0.5) minutes additional response time | <0.001 |
|  | Ambulance response time Category 2 (OM1b) | 12.3 (12.0, 12.7) minutes additional response time | <0.001 |
|  | Ambulance response time Category 3 (OM1c) | 38.7 (36.8, 40.5) minutes additional response time | <0.001 |
|  | Ambulance handover time (OM2) | 20.1 (19.7, 20.5) minutes additional handover time | <0.001 |
|  | Time until ED treatment for non-admitted patients (OM3) | 4.5 (4.1, 4.9) minutes additional time until treatment | <0.001 |
| +4 hours boarding time for medical patients | Inpatient LOS (OM4) | 13.0 (11.0, 14.9) hours additional LOS | <0.001 |
|  | 30-day readmission rate (OM5) | 2.9 (0.1, 5.1)% increase in odds of readmission within 30 days | <0.05 |
|  | 30-day mortality rate (OM6) | 6.4 (3.5, 9.4)% increase in odds of mortality within 30 days | <0.01 |

For the indirect/upstream impacts (OM1-3), most outcomes were underperforming against the national targets [24] (which is not uncommon in the NHS), with even the largest reductions in medicine patient boarding expected only to bring ambulance Category 1 response times within target (Figure 1). Here, while the effect size is relatively small, achieving below-75% boarding is sufficient to ensure the target is met. The effect is much more pronounced for Category 2 and 3 calls, whereby an extra one-fifth of ED bedded capacity used by medical boarders is estimated to add 12 and 39 minutes to ambulance response times respectively, and an additional 20 minutes for ambulance handover times. The effect on ED throughput is that when the number of medical boarding patients is less than half the number of majors and resus cubicles, 4 hour performance for non-admitted patients is 72.1%, whereas when there are more medical boarders than 50% of cubicles, only 66.9% of non-admitted are discharged within 4 hours.

**Figure 1.**
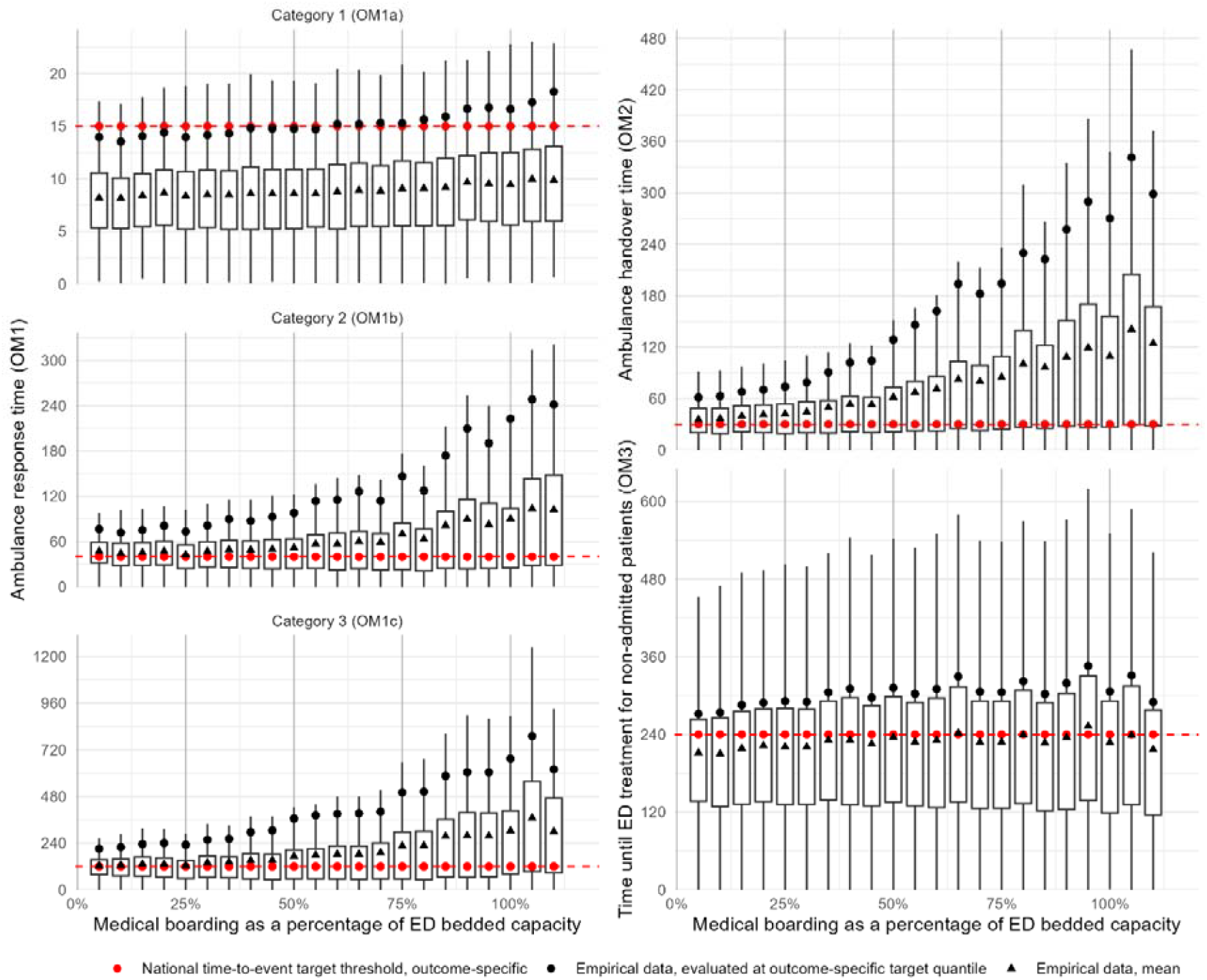
For indirect/upstream impacts (OM1-3), boxplots describing the distribution of outcome values (standardised for confounders and all in minutes) for different levels of medical boarding, as a percentage of ED bedded capacity. The box height represents the interquartile range (IQR), and whiskers are +/-1.5IQR.

For the direct/downstream impacts (OM4-6), results show that for every one-hour admission delay, inpatient LOS increases by just over three hours, such that a patient experiencing a boarding time of 24 hours will expect to spend 3.25 days longer on the wards (approximately 50% more LOS) than a patient who has experienced no admission delay whatsoever (Figure 2). While approximately a third of the value in absolute terms, the relative effect of boarding time on mortality is over double that of readmission risk. Accordingly, the risk of death within 30 days of admission increases from 3.5% to 5% for a patient experiencing 24 hours boarding time compared to one whose admission was not delayed.

**Figure 2.**
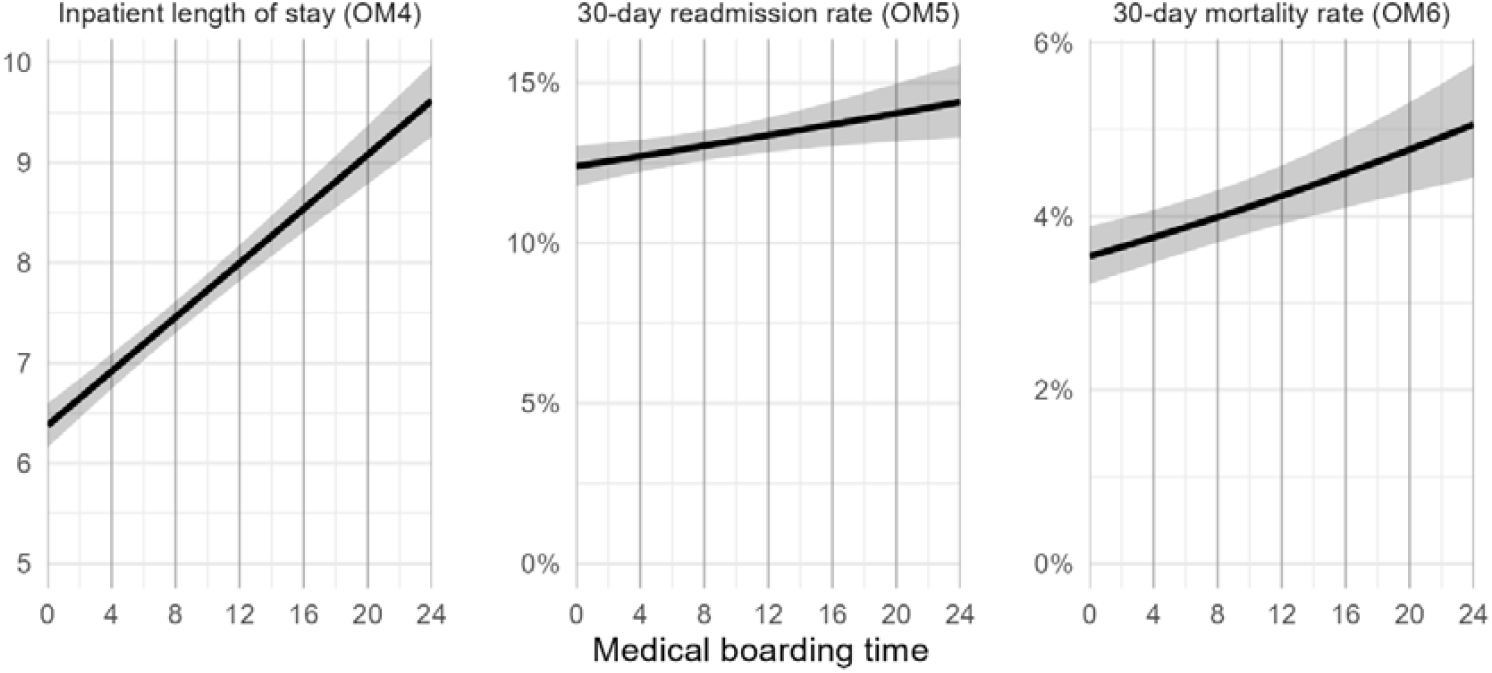
For direct/downstream impacts (OM4-6), predictor effect displays describing the mean (solid line) and 95% confidence intervals on the mean (shaded areas) for the outcome values based on different durations of medical boarding time (in hours). Note that inpatient length of stay (OM4) is measured in days.

## 4. Discussion

This study contributes to the existing literature by quantifying the impacts of medical patient boarding on the wider emergency patient pathway. The six outcome measures demonstrate how medical patient boarding may have significant adverse consequences for the outcomes of patients both directly affected, on medical wards following the ED, and those indirectly affected, awaiting ambulances and within the ED. Regression analysis has shown that for a typical 25-bed ED, each additional five medical boarders was associated with an extra 12 and 39 minutes for Category 2 and 3 ambulance response times (p<0.001) and an extra 20 minutes for ambulance handover times (p<0.001). For admitted medical patients, each additional 4 hours of boarding time was associated with an extra 13 hours inpatient length of stay (p<0.001) and a 6% increase in odds of 30-day mortality (p<0.01).

This study is new in that it is focused solely on the impact of medical patients boarding in the ED and the disproportionate impact on and of this group. This study has also standardised for confounding factors and collects data from 3 neighbouring but different types of hospital making results more generalisable. The outcomes measured show the impact of boarding on the medical patients themselves and also on other patients in the ED and those waiting for ambulances.

This is not a randomised controlled trial, and while correlation does not prove causation, the link from correlation to causation is supported by having controlled for potential sources of bias, in regard of demographic, socioeconomic, and clinical characteristics, healthcare activity attributes, and general performance levels (admission diagnosis and investigation data were considered but not included on account of the possible influence of boarding on these variables). Effect sizes are mostly relatively large, and all reported results are statistically significant to at least the 0.05 level. In regard of other Bradford Hill criteria [28], taking the impact on inpatient length of stay as an example: the association is strong with 13.0 (11.0 – 14.9) hours additional inpatient LOS for every 4 hours of boarding (p<0.001); the exposure clearly precedes the outcome; there is biological plausibility – typically older and frail medical patients waiting a long time for a medical bed, possibly in a non-cubicle escalation area, may miss medications, meals, and sleep, and so may decondition; and the finding is consistent with other studies showing increased LOS following boarding delays for hip fracture and ICU patients [29,30].

As well as leading to negative patient outcomes in the more immediate term, boarding may initiate a vicious cycle of delay related harm, which reduces inpatient bed availability and makes boarding in the ED more likely in subsequent days. Longer inpatient LOS for boarding patients (OM4) restricts the effective throughput of the hospital wards, thus reducing the chances of finding a medical bed for new ED patients requiring admission. At the same time, increased readmissions (OM5) place further demands on the inpatient wards, additionally constraining availability and so increasing the likelihood of boarding in the ED. It is assumed, often without contest, that the saturation of acute hospitals is due to an insufficient number of beds or lack of social care capacity. While these factors are likely to contribute, the results of this study suggest that part of the problem is the knock-on effects of boarding in the ED: reduce this and alleviate pressure on the acute hospital bed base, potentially limiting the need to outlie patients and easing the limitations on elective work. There could be much scope for improvement here: the mean 384-minute boarding time observed at the three hospitals of this study corresponds to an additional 21 hours of inpatient LOS for a typical medical patient admitted through the ED. Practically, the current thresholds to implement escalation measures within acute hospitals may be set too high, with long delays normalised. Avoiding this cycle of delay related harm would require greater discipline and a shift in management practices from reactively intervening to periods of prolonged boarding to proactively keeping boarding to a minimum at all times.

Future research could study the impact of ambulance handover and response times on mortality and inpatient length of stay. Further studies into the impact of ED boarding could examine whether increased boarding leads to increased social care requirements on discharge form hospital – either at home or in the level of residential care. While the study setting should be representative of the country at large – the three hospitals include a major trauma centre, a city centre teaching hospital, and a district general hospital in a smaller town – a larger study would nonetheless help validate that results hold at a national level. Finally, at a practical level, the models developed here could be implemented with routine ED data to estimate the (e.g.) weekly totals of boarding-induced additional ambulance response/handover times, ED/inpatient LOS, and readmissions/mortality. Using this data would enable a focus on quality improvement for both patient outcomes and system efficiency.

## Data Availability

All data produced in the present work are contained in the manuscript and supplementary material.

## Supplementary Material

**Table SM1.**
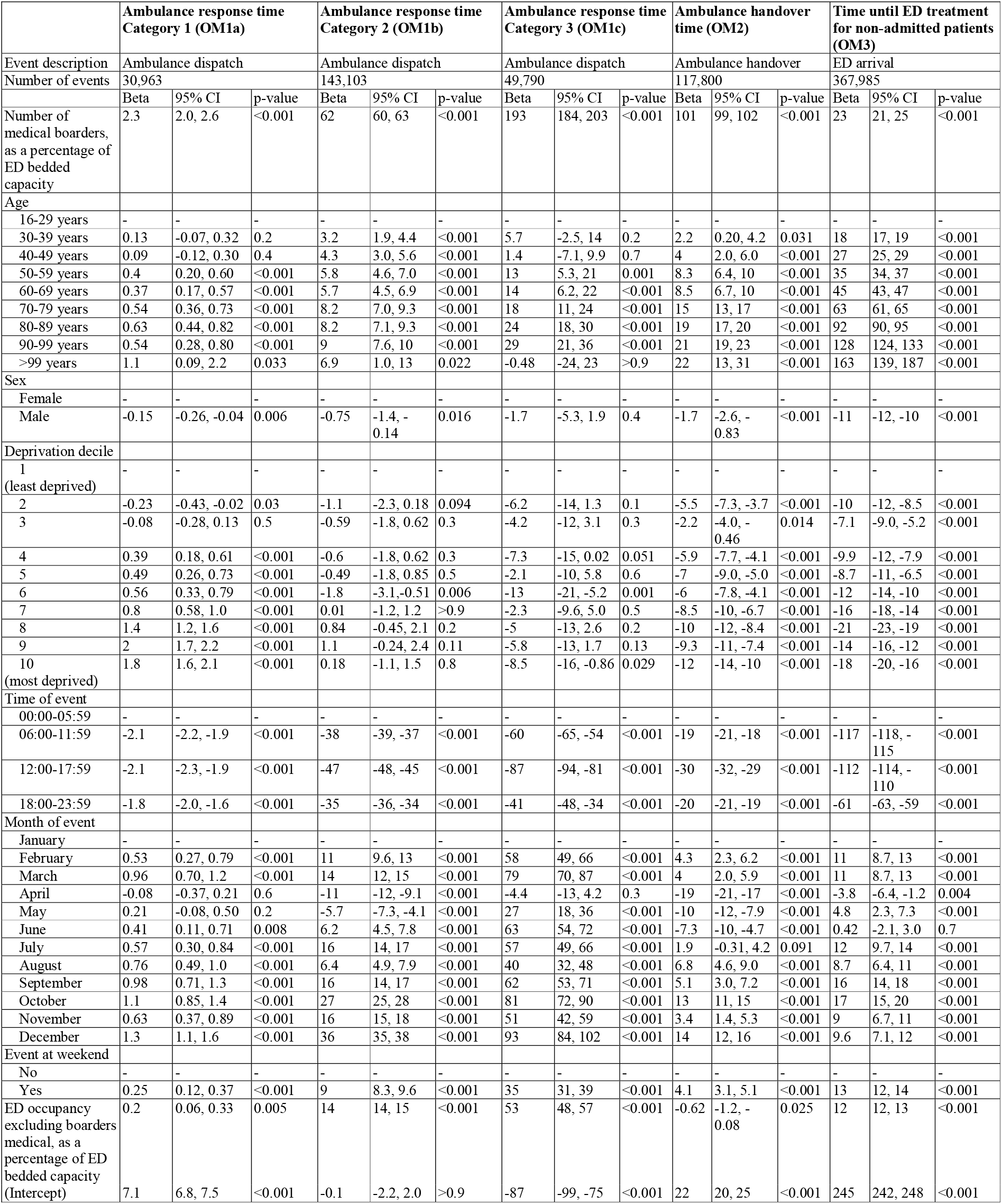
For indirect/upstream impacts (OM1-3), regression model parameters (including 95% confidence intervals) and accompanying p-values. Units are minutes.

**Table SM2.**
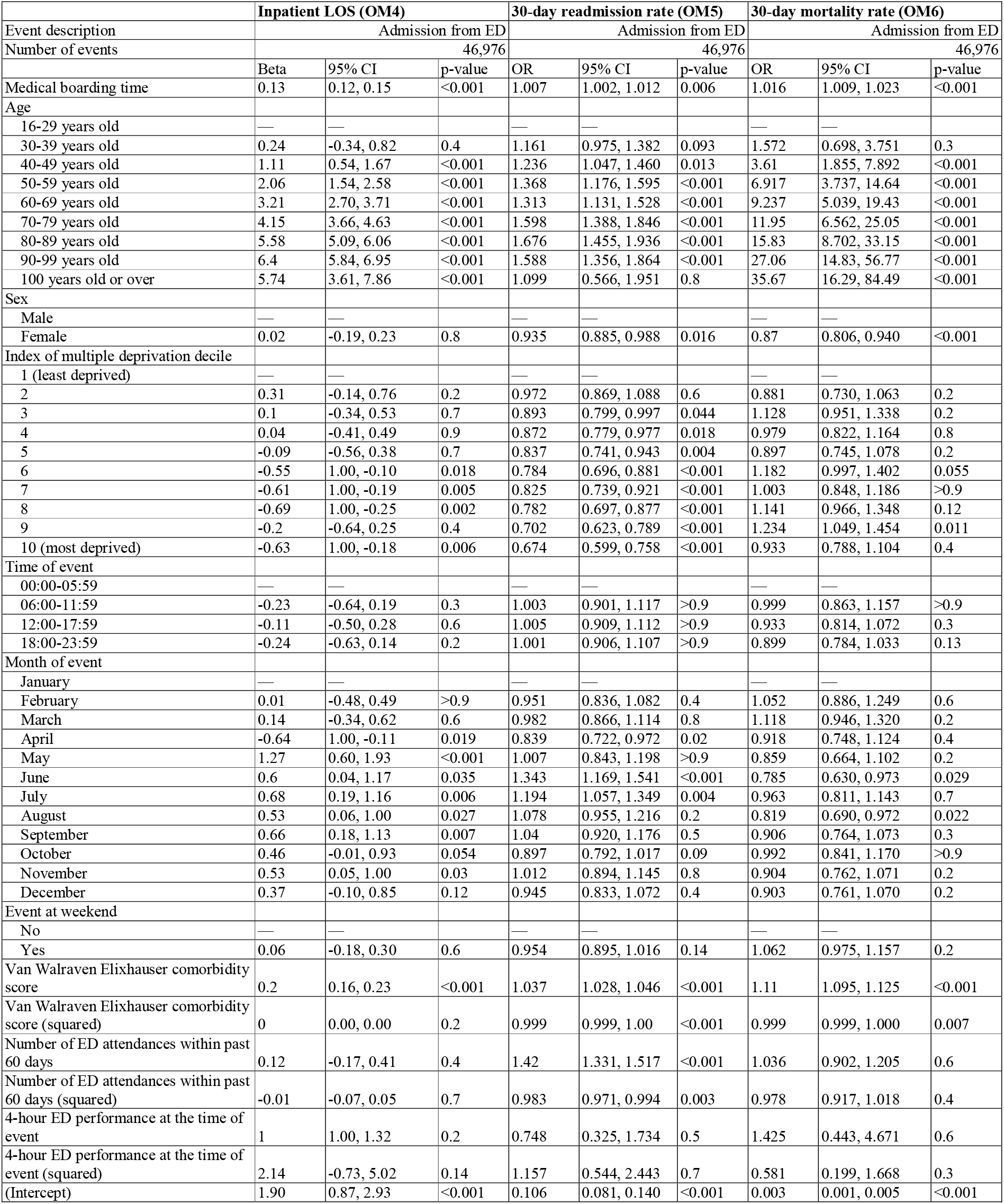
For direct/downstream impacts (OM4-6), regression model parameters (including 95% confidence intervals) and accompanying p-values. Units are hours.

